# Decreased Stroke, Acute Coronary Syndrome, and Corresponding Interventions at 65 US Hospitals Following COVID-19

**DOI:** 10.1101/2020.05.07.20083386

**Authors:** Adam de Havenon, John P. Ney, Brian Callaghan, Alen Delic, Samuel Hohmann, Ernie Shippey, Shadi Yaghi, Mohammad Anadani, Gregory J. Esper, Jennifer J. Majersik

## Abstract

**Background:** Following the emergence of coronavirus disease 2019 (COVID-19), early reports suggested a decrease in stroke and acute coronary syndrome (ACS). We sought to provide descriptive statistics for stroke and ACS from a sample of hospitals throughout the United States, comparing data from March 2020 to similar months pre-COVID.

**Methods:** We performed a retrospective analysis of 65 academic and community hospitals in the Vizient Clinical Data Base. The primary outcome is monthly count of stroke and ACS, and acute procedures for both, from February and March in 2020 compared to the same months in 2018 and 2019. Results are aggregated for all hospitals and reported by Census Region.

**Results:** We identified 51,246 strokes (42,780 ischemic, 8,466 hemorrhagic), 1,043 mechanical thrombectomies (MT), 836 tissue plasminogen activator (tPA) administrations, 36,551 ACS, and 3,925 percutaneous coronary interventions (PCI) for ACS. In February 2020, relative to February 2018 and 2019, hospitalizations with any discharge diagnosis of stroke and ACS increased by 9.8% and 12.1%, respectively, while in March 2020 they decreased 18.5% and 7.5%, relative to March 2018 and 2019. When only including hospitalizations with the primary discharge diagnosis of stroke or ACS, in March 2020 they decreased 17.6% and 25.7%, respectively. In March 2020, tPA decreased 3.3%, MT increased 18.8%, although in February 2020 it had increased 36.8%, and PCI decreased 14.7%. These decreases were observed in all Census regions.

**Conclusions:** Following greater recognition of the risks of COVID-19, hospitalizations with stroke and ACS were markedly diminished in a geographically diverse sample of United States hospitals. Because the most likely explanation is that some patients with stroke and ACS did not seek medical care, the underlying reasons for this decrease warrant additional study to inform public health efforts and clinical care during this and future pandemics.

## Introduction

After the emergence of coronavirus disease 2019 (COVID-19), there have been early reports of a paradoxical effect on stroke and acute coronary syndrome (ACS). Instead of hospitalization rates remaining stable or increasing, which was expected because viral infections are a risk factor for stroke and ACS,^1,2^ their rate may have decreased. For stroke, single center reports and sharing of national experiences also suggests a marked decrease in stroke admissions.^3,4^ In Italy, the outbreak of COVID-19 was associated with a 32% decline in the number of percutaneous coronary interventions (PCI) for ACS.^5^ A study of 7 centers in the United States reported a 38% decrease in cardiac catheterization laboratory activations in March 2020.^6^ To inform clinical care and public health during this and future pandemics, it is crucial to provide reliable estimates of this decrease.

## Methods

We performed a retrospective analysis using the Vizient^®^ Clinical Data Base (CDB), a health care analytics platform.^7^ The analysis was of de-identified data at the hospital level and included hospitals with data from February and March 2018–20, which restricted us to “early reporting” hospitals in March 2020. The de-identified data did not necessitate IRB approval and is available upon reasonable demand from Vizient.

We identified five outcomes through discharge diagnosis and billing codes in any position: 1) stroke, comprised of acute ischemic stroke (AIS) and hemorrhagic stroke (intracerebral and subarachnoid hemorrhage; 2) mechanical thrombectomy (MT) for AIS; 3) tissue plasminogen activator (tPA) administration for AIS; 4) ACS, comprised of acute myocardial infarction, unstable angina, and other acute ischemic heart disease; and 5) PCI for ACS. The codes used for identification are found in Supplemental Table 1. We performed a second analysis where we only included cases with stroke and ACS discharge diagnosis codes in the primary position.

**Table 1.**
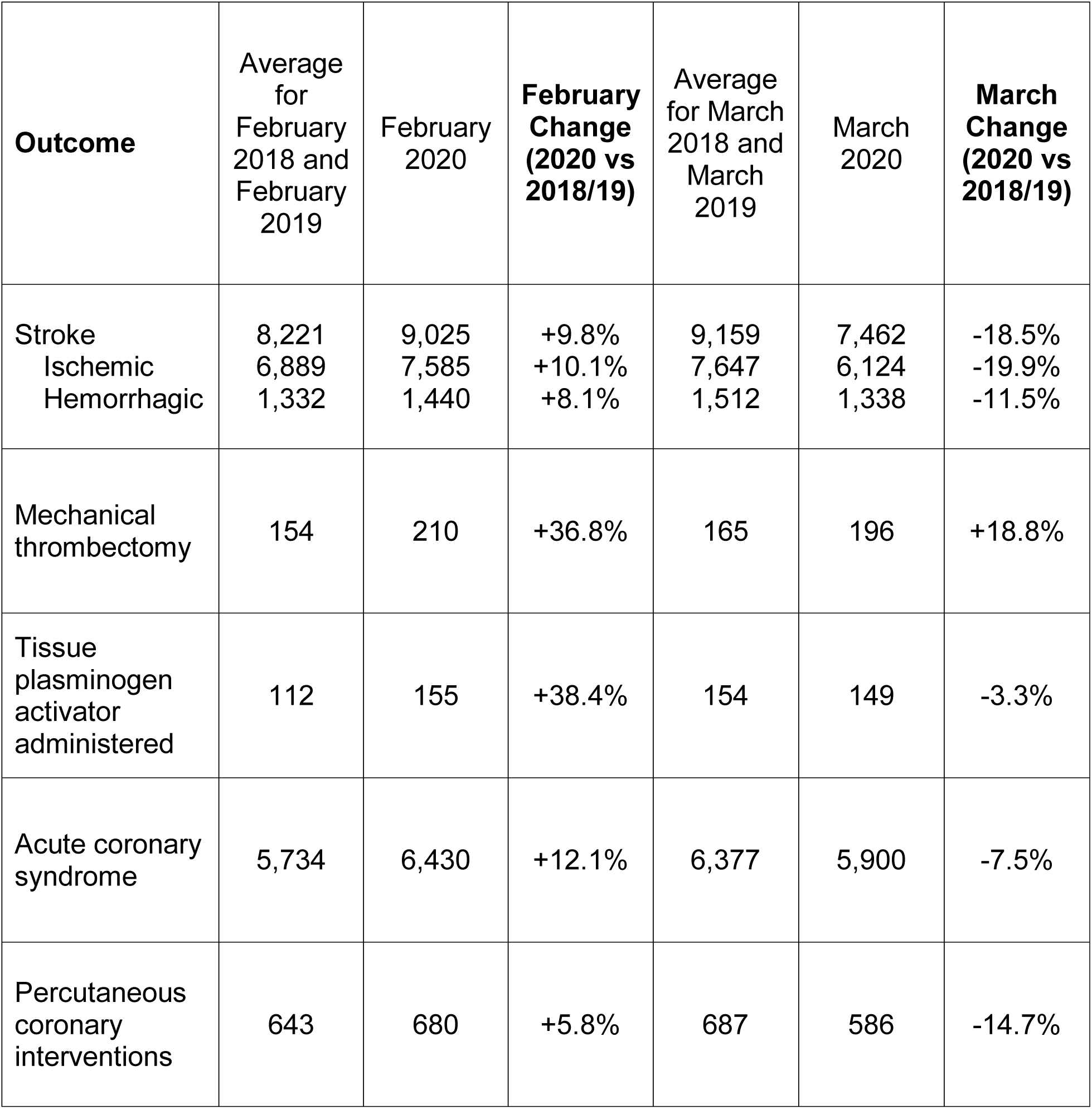
Hospitalizations with acute coronary syndrome and stroke, and procedures for both, for 65 hospitals in February and March 2020 versus the average of 2018 and 2019.

| **Outcome** | Average for February 2018 and February 2019 | February 2020 | **February Change (2020 vs 2018/19)** | Average for March 2018 and March 2019 | March 2020 | **March Change (2020 vs 2018/19)** |
| --- | --- | --- | --- | --- | --- | --- |
| Stroke | 8,221 | 9,025 | +9.8% | 9,159 | 7,462 | -18.5% |
| Ischemic | 6,889 | 7,585 | +10.1% | 7,647 | 6,124 | -19.9% |
| Hemorrhagic | 1,332 | 1,440 | +8.1% | 1,512 | 1,338 | -11.5% |
| Mechanical thrombectomy | 154 | 210 | +36.8% | 165 | 196 | +18.8% |
| Tissue plasminogen activator administered | 112 | 155 | +38.4% | 154 | 149 | -3.3% |
| Acute coronary syndrome | 5,734 | 6,430 | +12.1% | 6,377 | 5,900 | -7.5% |
| Percutaneous coronary interventions | 643 | 680 | +5.8% | 687 | 586 | -14.7% |

We report descriptive statistics for February and March in all years, compare the outcome counts in 2020 to the monthly averages from 2018 and 2019, and report the within year percent change from February to March. To provide a geographic distribution, we describe the hospital location by United States Census region.^8^

## Results

We included data from 65 hospitals, of which 55 were academic medical centers and 20 had less than 200 beds, 14 had 200–499 beds, and 21 had ≥500 beds. There were 17 hospitals in the South, 6 in the Northeast, 11 in the West, and 31 in the Midwest. We identified 51,246 hospitalizations with stroke (42,780 AIS, 8,466 hemorrhagic), 1,043 MTs for AIS, 836 tPA administrations for AIS, 36,551 hospitalizations with ACS, 3,925 PCIs for ACS. In February 2020, compared to the average of February 2018 and 2019, there was an increase in all outcomes.

In March 2020, compared to the average of March 2018 and 2019, there were decreases in stroke (18.5%), tPA (3.3%), ACS (7.5%), and PCI (14.7%) (Table 1, Figure 1). There was an 18.8% increase in MT, but in February 2020 the increase was 36.8%. Similar results were seen in the secondary analysis restricted to patients with stroke or ACS in the primary diagnosis position (Table 2, Figure 2), which showed a March 2020 decrease of 17.6% for stroke and 25.7% for ACS. In that analysis, acute ischemic stroke decreased 17.9% and hemorrhagic stroke decreased 15.7%.

**Figure 1.**
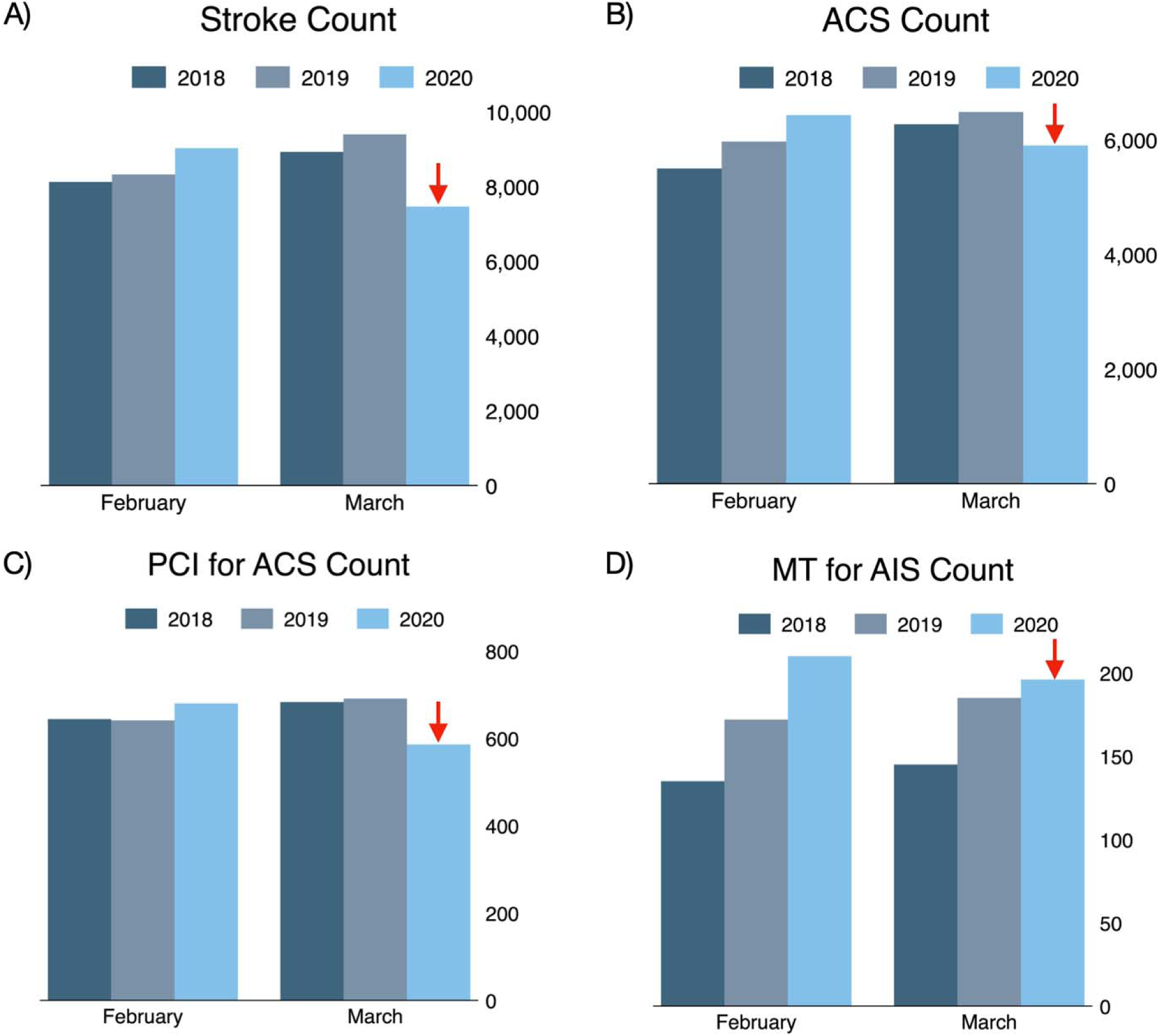
Monthly counts of outcomes for February and March 2018–2020, shown for all all 65 hospitals, with March 2020 indicated by a red arrow. A) stroke, B) acute coronary syndrome (ACS), C) mechanical thrombectomy (MT) for AIS, and D) percutaneous coronary intervention (PCI) for ACS.

**Table 2.**
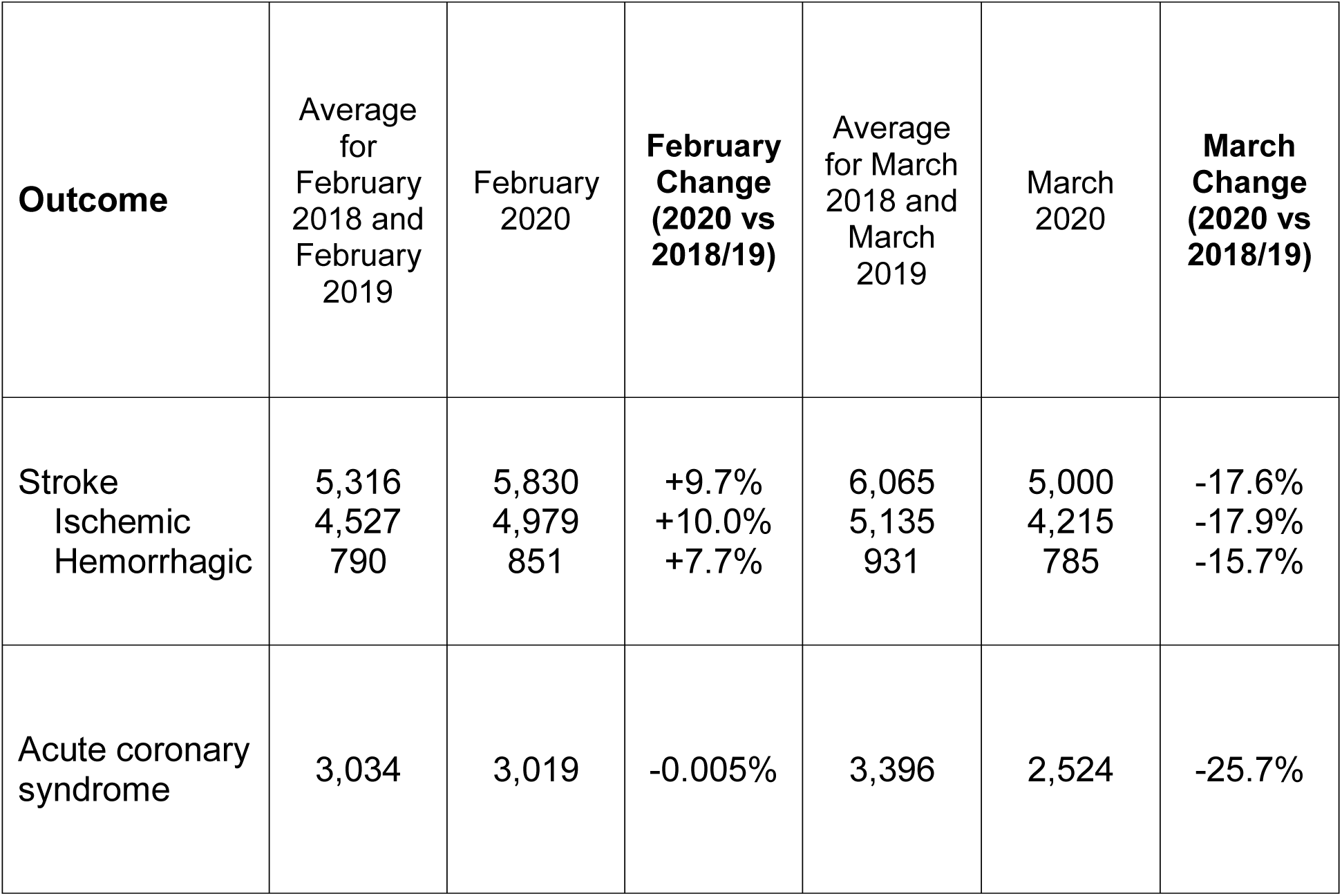
Cases that had the primary discharge diagnosis codes for stroke and acute coronary syndrome at 65 hospitals in February and March 2020 versus the average of 2018 and 2019.

| **Outcome** | Average<br>for<br>February<br>2018 and<br>February<br>2019 | February<br>2020 | **February**<br>**Change**<br>**(2020 vs**<br>**2018/19)** | Average<br>for March<br>2018 and<br>March<br>2019 | March<br>2020 | **March**<br>**Change**<br>**(2020 vs**<br>**2018/19)** |
| --- | --- | --- | --- | --- | --- | --- |
| Stroke | 5,316 | 5,830 | +9.7% | 6,065 | 5,000 | -17.6% |
| Ischemic | 4,527 | 4,979 | +10.0% | 5,135 | 4,215 | -17.9% |
| Hemorrhagic | 790 | 851 | +7.7% | 931 | 785 | -15.7% |
| Acute coronary<br>syndrome | 3,034 | 3,019 | -0.005% | 3,396 | 2,524 | -25.7% |

**Figure 2.**
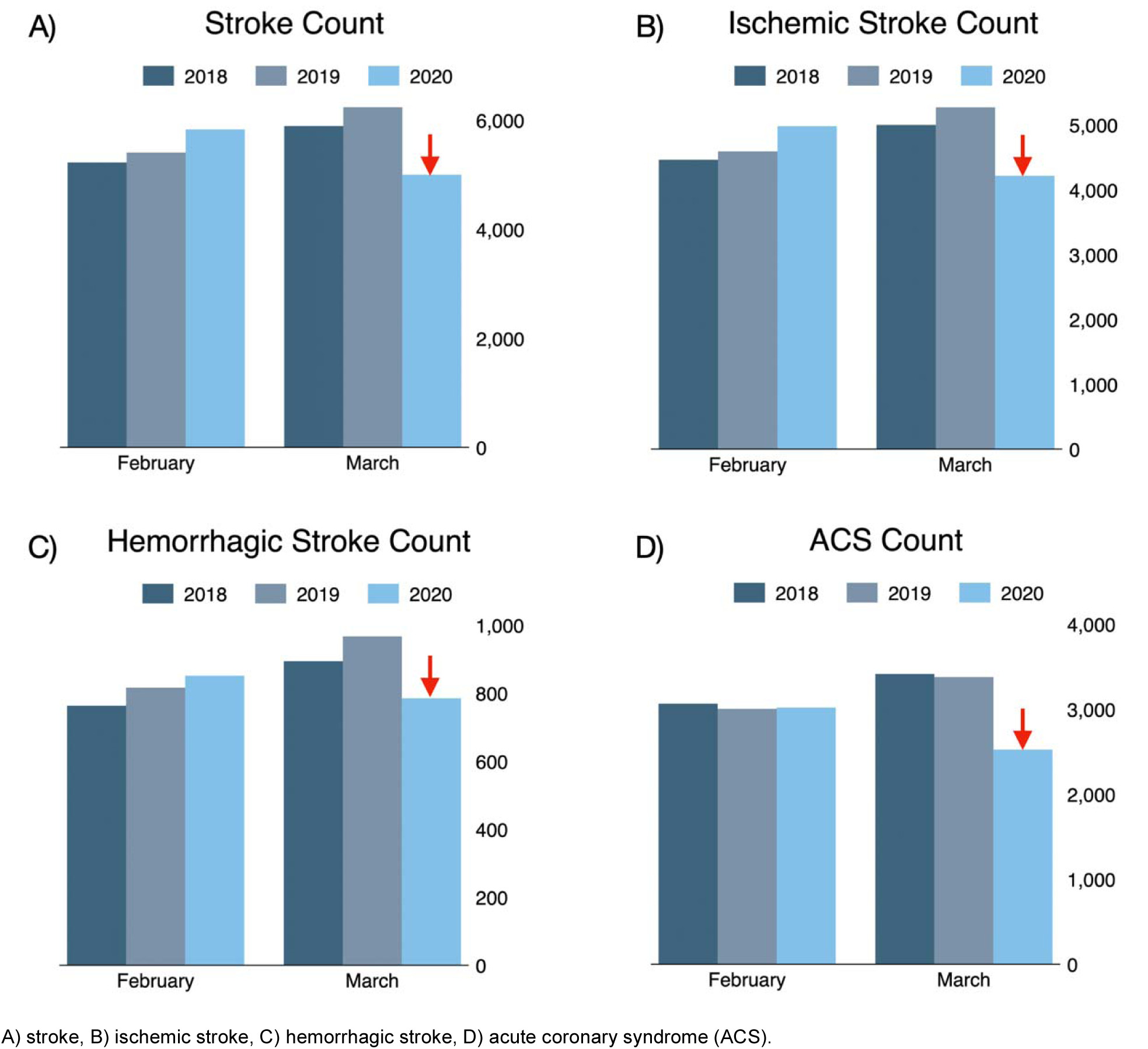
Monthly counts for cases that had the primary discharge diagnosis codes for acute coronary syndrome and stroke for February and March 2018–2020, shown for all 65 hospitals.

We compared the percent change from February to March by Census region and saw that while in 2018 and 2019 there was an increase in our outcomes, in 2020 there was a comparative and absolute decrease (Supplemental Figure 1). There were regional differences, but the overall trend was decreases throughout the United States. For example, compared to the March average in 2018 and 2019, stroke decreased in March 2020 by 21.4% in the Midwest, 17.6% in the Northeast, 10.4% in the South, and 22.7% in the West. Additional regional comparisons are shown in Table 3. One outlier was a March 2020 increase in ACS of 18.4% in the Northeast, compared to decreases in all other Census regions.

**Table 3.** Hospitalizations with acute coronary syndrome and stroke, and procedures for both, at 65 hospitals in February and March 2020 versus the average of 2018 and 2019, by United States Census regions.

| **Outcome** | **Average for February 2018 and February 2019** | **February 2020** | **February Change (2020 vs 2018/19)** | **Average for March 2018 and March 2019** | **March 2020** | **March Change (2020 vs 2018/19)** |
| --- | --- | --- | --- | --- | --- | --- |
| Stroke, ischemic and hemorrhagic |  |  |  |  |  |  |
| Midwest | 3,033 | 3,387 | +11.7% | 3,431 | 2,696 | -21.4% |
| Northeast | 1,237 | 1,424 | +15.1% | 1,352 | 1,114 | -17.6% |
| South | 1,954 | 2,100 | +7.5% | 2,189 | 1,960 | -10.4% |
| West | 1,997 | 2,114 | +5.9% | 2,188 | 1,692 | -22.7% |
| Mechanical thrombectomy or tPA administered |  |  |  |  |  |  |
| Midwest | 88 | 138 | +56.8% | 110 | 92 | -16.0% |
| Northeast | 36 | 57 | +58.3% | 46 | 58 | +26.1% |
| South | 96 | 117 | +21.9% | 115 | 130 | +13.0% |
| West | 46 | 53 | +16.5% | 49 | 65 | +34.0% |
| Acute coronary syndrome |  |  |  |  |  |  |
| Midwest | 2,303 | 2,579 | +12.0% | 2,632 | 2,391 | -9.2% |
| Northeast | 762 | 958 | +25.7% | 809 | 958 | +18.4% |
| South | 1,348 | 1,465 | +8.7% | 1,459 | 1,347 | -7.7% |
| West | 1,322 | 1,428 | +8.0% | 1,478 | 1,204 | -18.5% |
| Percutaneous coronary intervention |  |  |  |  |  |  |
| Midwest | 259 | 276 | +6.8% | 267 | 234 | -12.2% |
| Northeast | 71 | 78 | +9.8% | 72 | 64 | -11.1% |
| South | 196 | 195 | -0.003% | 197 | 163 | -17.1% |
| West | 118 | 131 | +11.5% | 152 | 125 | -17.8% |

## Discussion

There was a substantial decrease in the number of patients discharged from hospitals across the United States with stroke and ACS in March 2020 relative to March of prior years and a decrease in tPA for AIS and PCI for ACS, but only a relative decrease in MT for AIS compared to the expected volume. This may be because new indications for MT in 2018 increased the number of eligible patients.^9^ In February 2020, MT for AIS increased 36.8%, while in March 2020 it only increased 18.8%, suggesting that MT volumes may also have been affected during the COVID-19 pandemic. In 2018 and 2019, there were higher rates of all outcomes in March compared to February of the same year, which is a previously reported phenomenon.^10–12^

The decrease in stroke and ACS in March 2020 corresponds to not only rising COVID-19 infections, but also widespread recognition of infection risk and accompanying restrictions in movement and business activity.^13^ In contrast, our analysis of February showed the opposite – a 2020 increase in stroke and ACS, consistent with overall increasing volumes at the academic hospitals that are well-represented in this sample. The change of pattern we observed suggests a disruptive cause that is not part of the historical stroke and ACS trends at these hospitals.

Infection with respiratory viruses confers a higher risk of stroke and ACS^1,2^ and COVID-19 infection is thought to be pro-thrombotic.^14^ Because COVID-19 may increase the incidence of stroke and ACS, the observed decrease suggests that other factors accounted for our findings. The most likely explanation is that patients with less severe stroke and ACS were not seeking medical care because of concern about acquiring COVID-19 infection. For stroke, this theory is supported by the relative preservation of MT, which is reserved for patients with more severe stroke. However, PCI for ACS, the cardiac analog of MT, decreased 14.7% in March 2020.

Other potential causal factors include that the reduction in automobile usage and furloughing of factories has improved air pollution,^15^ which is a well-established short-term and long-term risk factor for stroke and ACS.^16^ Additionally, medication adherence or wellness may have improved during quarantine and the decline in elective procedures may have reduced peri-procedural stroke and ACS. Lastly, the reduction could be due to an unknown protective effect of COVID-19 infection.

Our study has several limitations, including that it is not a fully representative sample and that case identification with administrative and billing codes has bias. For example, we captured less MT and tPA cases compared to expected rates,^17^ but the methodology and sample was consistent across 6 time points, lending it validity. With the current data we are not able to explore the causal factors for the reduction and this phenomenon may be part of a greater year over year trend that is not captured in three years of data. Despite these limitations, we have taken a critical step by showing that the reduction exists beyond single centers and across Census regions. Future work will confirm these findings in larger datasets and analyze potential mediators.

## Conclusion

In March 2020, after the rise in COVID-19 infections and accompanying restrictions in movement and business activity in the United States, the rate of stroke, ACS, and corresponding interventions at hospitals across the country declined. This decrease is important given the high morbidity, mortality, and efficacious interventions for stroke and ACS. Because the most likely explanation is that some patients with stroke and ACS were not seeking medical care, public health messaging should include reminders that emergent care for acute cardiovascular and neurologic symptoms should not be postponed during a pandemic. As restrictions of some form may remain in place for the near future, the definitive reasons for this decrease warrant additional study to inform public health efforts and clinical care during this and future pandemics.

## Data Availability

The data is not available.

## Acknowledgements

None

## Sources of Funding

Dr. de Havenon is supported by NIH-NINDS K23NS105924.

## Disclosures

Dr. de Havenon has received investigator initiated funding from AMAG and Regeneron pharmaceuticals. Dr. Callaghan consults for a PCORI grant, DynaMed, and performs medical legal consultations including consultations for the Vaccine Injury Compensation Program. Dr. Majersik reports NIH/NINDS funding U24NS107228, funding for Associate Editor at *Stroke*, Consulting fees for Foldax scientific advisory board, and is on Editorial Board member of *Neurology*. Dr. Esper reports consult fees from NeuroOne Medical Technologies, Inc and fees as a medicolegal consultation expert witness. The remaining authors report no potential conflicts of interest.

## Notes

### Competing Interest Statement

The authors have declared no competing interest.

